# Doubtful Clinical Benefit of Casirivimab-Imdevimab Treatment for Disease Severity Outcome of High-Risk Patients with SARS-CoV-2 Delta Variant Infection

**DOI:** 10.1101/2022.01.29.22270090

**Authors:** Noah Shopen, Michal Dekel, Michal Mizrahi, Efrat Zandberg, Nancy Bishouty, Daniel Talmud, Ben Vaknin, Shira Haberman, Malka Katz Shalhav, David Zeltser, Neta Cohen

## Abstract

Casirivimab/Imdevimab therapy reportedly retains neutralization potency against circulating SARS-CoV-2 variants, including Delta (B.1.617.2), but there are sparse data on its clinical benefit against the Delta variant among vaccinated and unvaccinated patients. We explored its therapeutic effect on COVID-19 severity outcome in terms of room air saturation <93% within 14 days of initial presentation and 45-day all-cause mortality among high-risk patients with SARS-CoV-2 Delta variant infection and compared its effect between vaccinated and unvaccinated patients. We conducted a retrospective cohort study at a tertiary care medical center between 6/2021 and 9/2021 and included patients who presented with a positive PCR for SARS-CoV-2 and fulfilled the criteria for Casirivimab/Imdevimab treatment. Of the 359 suitable patients (52% female, median age 63 years), 116 were treated with Casirivimab/Imdevimab and 243 were not. Two-hundred and one (56%) patients had received at least 2 SARS-CoV-2 vaccinations. Casirivimab/Imdevimab treatment was not an independent protective factor of COVID-19 severity outcome (multivariable analysis). Chronic kidney disease (aOR=3.51 [95%CI: 1.34-9.20], *P*=0.01), lower saturation levels (aOR=0.7 [95%CI: 0.58–0.85], *P*<0.01), abnormal chest x-ray findings (aOR=2.92, [95%CI: 1.24–6.87, *P=*0.01), and higher C-reactive protein levels (aOR=1.01 [95%CI: 1.00–1.01], *P*=0.008) were independent risk factors of COVID-19 severity. Positive immunization status was an independent protective factor (aOR=0.33 [95%CI: 0.14–0.77], *P*=0.01). A sub analysis of patients treated with Casirivimab/Imdevimab revealed no significant difference in COVID-19 severity between vaccinated and unvaccinated patients. These findings demonstrate no added benefit of Casrivimab/Imdevinab treatment for high-risk patients with the SARS-CoV-2 Delta variant infection, regardless of their vaccination status.

## INTRODUCTION

The ongoing COVID-19 pandemic continues to be a global threat. Up to 10% of patients initially presenting with mild illness eventually progress to a severe disease (1). Most of those patients have a least one comorbid condition (2). Treatment with neutralizing monoclonal antibodies (mAbs) for COVID-19 is based upon several randomized placebo-controlled clinical trials that led to emergency use authorization (EUA) by the US Food and Drug Administration (FDA) (3-5). REGEN-COV, a combination of 2 neutralizing monoclonal antibodies, Casirivimab and Imdevimab, binds noncompeting epitopes of the receptor-binding domain of the SARS-CoVID-19 spike protein (6), and has been shown to rapidly reduce the viral load, shorten the duration of symptoms, and reduce the need for hospitalization and the risk of death in high-risk non-hospitalized patients with COVID-19 (7-9). Delta variant became a variant of concern by WHO on May 11, 2021 (10). While Casirivimab/Imdevimab therapy was found to retain neutralization potency against circulating SARS-CoV-2 variants of concern, including delta (B.1.617.2) in vitro and in vivo (6,11,12), there are only sparse data (13) on its clinical benefit against the Delta variant.

A recent study on patients infected with non-delta SARS-CoV-2 found that Casirivimab/Imdevimab reduced the 28-day mortality among hospitalized patients who were seronegative at baseline (14). Most of the other relevant studies had been performed prior to the vaccination era, and little has been published with respect to its effect among vaccinated patients (13). An exception is Bierle et al (13) who found monoclonal antibody treatment to be associated with reduced hospitalization in vaccinated high-risk persons with mild-to moderate COVID-19. However, and their study was initiated prior to May 2021, it was not focused on the Delta variant.

We aimed to determine the effect of monoclonal antibody Casirivimab/Imdevimab treatment on SARS-CoV-2 disease severity defined as saturation <93% within 14 day of initial presentation and 45-day all-cause mortality, among high-risk patients with a SARS-CoV-2 Delta variant infection, and to compare that effect between vaccinated and unvaccinated patients.

## MATERIALS AND METHODS

### Ethics

Approval for this study was obtained from our institutional research ethics board. The study meets the STROBE criteria and is reported according to the STROBE guideline (15).

### Study design and population

We conducted a retrospective cohort study at the emergency department (ED) of a tertiary university-affiliated medical center between June 1, 2021 and September 31, 2021. Included were all patients who presented with a positive PCR for SARS-CoV-2 to the ED or were identified as being positive while hospitalized for a non-COVID-19 indication. All of the study participants fulfilled the criteria for receiving Casirivimab/Imdevimab treatment (see below).

### Inclusion and exclusion criteria

The inclusion criteria were based upon the FDA (EUA) guidelines (16). Patients who presented to our hospital during the study period and fulfilled all of the following criteria:

1. A positive PCR for SAR-COV-2 within 10 days of presentation.
2. High-risk factors for developing severe COVID-19 infection, defined as one of the following criteria (16): age >65 years, age >55 years with a chronic heart condition (including ischemic heart disease, chronic dysrhythmias, heart failure, and valves and structural illness) or a pulmonary disease (defined as chronic obstructive pulmonary disease or heavy smoking over 10 pack years), and older than 12 years of age with any of the following: diabetes mellitus, chronic kidney disease, morbid obesity (defined as body mass index ≥35), being severely immunocompromised (organ transplant patients, bone marrow transplant patients, hematologic malignancy, patients receiving anti-CD20 or Fingolimod, congenital immunodeficiency and acquired immunodeficiency, including HIV with a CD4 count <300), pregnancy patients and those with liver failure.

Patients who were diagnosed with severe COVID-19 infection already at initial presentation (room air saturation <93%) were excluded.

### Data collection

The parameters that were extracted from the medical records included date of visit, age, sex, medical history, immunization status, time from symptom onset (days), time from diagnosis (days), symptoms, and vital signs upon arrival to the ED (room air saturation, pulse, blood pressure, body temperature). In addition, we collected laboratory and imaging test results, including white blood cell count and differential, neutrophil/lymphocytes ratio, C–reactive protein (CRP) level, SARS-CoV-2 IgG antibodies titers (in U/ml), and abnormal chest x-ray findings (see below). Treatments and medications were documented, including implementation of an O2 cannula, non-invasive high flow respiratory support system, as well as invasive ventilation, antibiotics, steroids, anticoagulants and Casirivimab/Imdevimab administration. Also noted were length of stay (in hours) in the ED and disposition (discharge/ward/intensive care unit), room air saturation <93% within 14 days of initial presentation, and 45-day all-cause mortality.

### Definitions

Vaccinated patients were defined as those who received at least 2 doses of the BNT162b2 vaccine, and unvaccinated patients were defined as those who received fewer than 2 doses. “Early” presentation was defined as a positive PCR for SAR-COV-2 result and symptoms onset within 10 days prior of arrival to the ED.

### Casirivimab/Imdevimab therapy and selection of recipients

Therapy consisted of a single parenteral injection of 1200 mg Casirivimab and 1200 mg Imdevimab. Patient selection was based upon the clinical decision of both the ED physician and the infection disease specialist, together with the patient’s agreement.

### Antibody titer measurement

Antibodies titers were measured with the ADVIA Centaur SARS-CoV-2 IgG (sCOVG) assay, which is designed for in vitro diagnostic use in the qualitative and quantitative detection of IgG antibodies, including neutralizing antibodies, to SARS-CoV-2 in human serum and plasma. The system reports sCOVG assay results in index values and as nonreactive or reactive: nonreactive was defined by an index of <1.0, and those samples were considered negative for SARS-CoV-2 IgG antibodies, while reactive was defined by an index of ≥1.0, and those samples were considered positive for SARS-CoV-2 IgG antibodies.

### Abnormal chest x-ray findings

Findings of consolidation, ground glass opacities, and nodules were interpreted as being abnormal. X-ray evidence of those occupying <50% of the lung field was defined as mild-to-moderate, and those occupying >50% of the lung field as severe (17).

### Outcome measurements

Our primary outcome was a composite of COVID-19 disease severity, whose definition was based upon previous studies (14,18,20) as either room air saturation <93% within 14 days of initial presentation or 45-day all-cause mortality. Given that hospitalization can be due to non-COVID-19 indications, we considered desaturation and mortality to be appropriate measurements of COVID-19 disease severity. Epidemiologic and clinical characteristics, as well as disease severity based on their Casirivimab/Imdevimab treatment were compared between the study patients. We then compared the occurrence of severe disease outcome between Casirivimab/Imdevimab vaccinated and unvaccinated patients and between seronegative and seropositive patients. Finally, epidemiologic, and clinical characteristics, including Casirivimab/Imdevimab treatment, were compared between patients with and without severe disease outcome. Only parameters that were with *P value* <0.15 in the univariable analysis or were considered as being clinically important (positive immunization status) were inserted into a multi-regression model to explore independent predictors for severe disease outcome.

### Statistical analysis

Data entry and analysis were performed with SPSS Statistics, version 26 (SPSS Inc, Chicago, IL). Categorical variables were described as number (percentage) and continuous variables as mean ± standard deviation, median, and range. Differences between 2 groups in continuous variables were assessed with the T-test, differences between categorical variables were assessed with the chi-square or Fisher exact test, and differences between medians were assessed with a Mann-Whitney U test for independent means. A binary logistic regression for severe disease outcome was performed to identify independent predictors for severe disease outcome. Parametric correlations were assessed by the Spearman coefficient. A *P*-value <0.05 was considered statistically significant.

## RESULTS

Three-hundred and fifty-nine patients were included in the final cohort. One-hundred and eighty-nine were females (52.6%), and the median age for the entire group was 63 years (IQR 41.0-75.0). Two-hundred and three patients (56.5%) received at least 2 doses of COVID-19 vaccine and 57 of them were vaccinated with a 3^rd^ dose. All of the patients fulfilled the inclusion criteria for Casirivimab/Imdevimab treatment eligibility: 116 of them were treated with Casirivimab/Imdevimab (“cases”) and 243 were not (“controls”) (Figure 1). Fifty of the 359 patients had a severe disease outcome, including 7 who died within 45 days from their initial presentation and 48 who had room air saturation <93% within 14 days from their initial presentation.

**FIG. 1.**
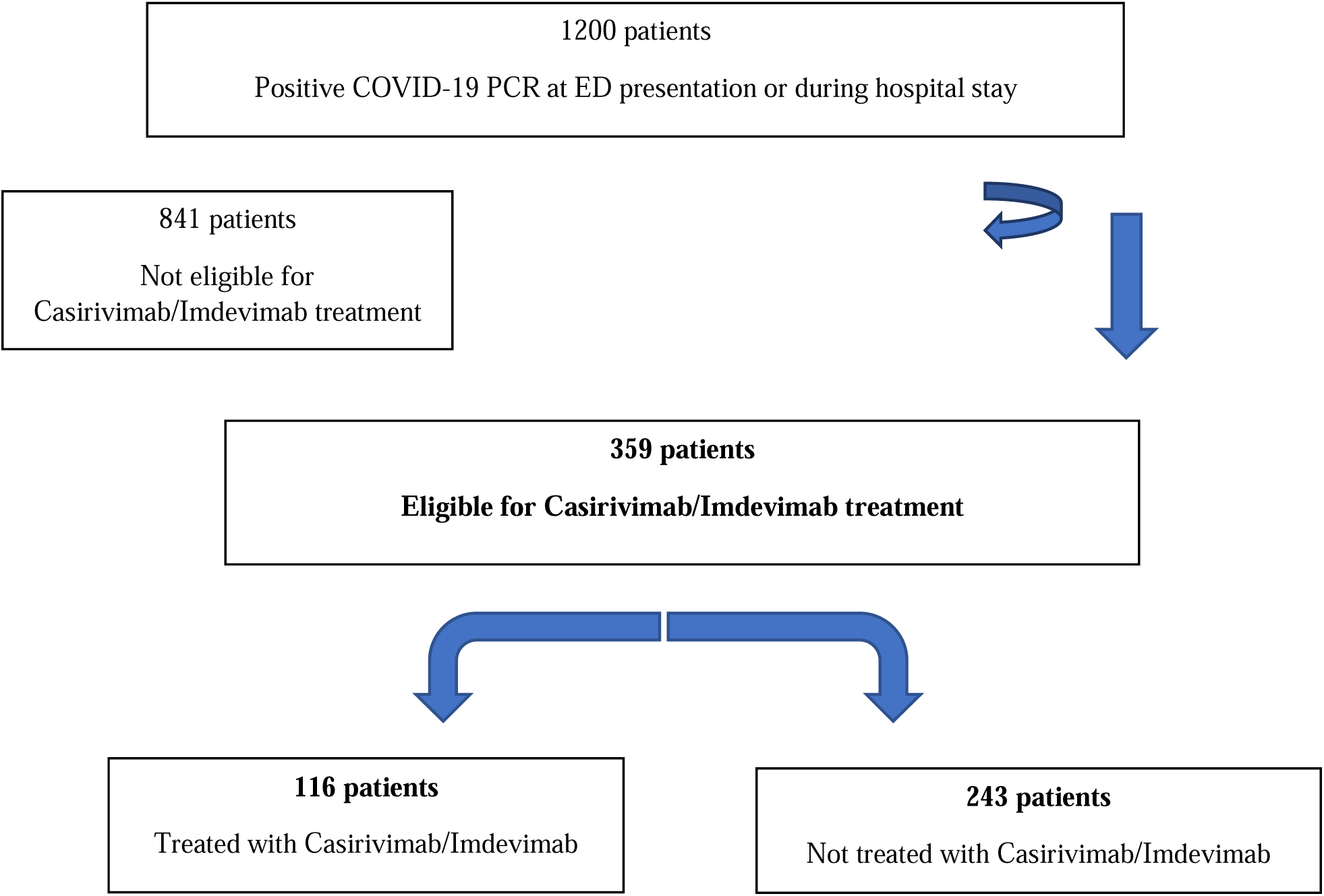
Flowchart of the study cohort

### Comparison of epidemiological and clinical characteristics, and severe disease outcome according to receiving or not Casirivimab/Imdevimab treatment

Table 1 displays the comparison of clinical and epidemiologic characteristics between patients who were treated with Casirivimab/Imdevimab and those who were not.

**TABLE 1.** Epidemiologic and clinical characteristics of Casirivimab/Imdevimab-treated and not treated patients

| Characteristic | Not treated<br>(n=243) | Treated<br>(n=116) | P value |
| --- | --- | --- | --- |
| Age, y (mean ± SD) | 57.6±22.2 | 62.2±18.3 | 0.03 |
| Sex (female, %) | 135 (55.2) | 55 (47.4) | 0.29 |
| Chronic disease: |  |  |  |
| Heart disease | 55 (22.6) | 34 (29.3) | 0.19 |
| Lung disease | 35 (14.5) | 24 (20.7) | 0.17 |
| Immunosuppression | 58 (24.0) | 56 (48.3) | <0.001 |
| Pregnancy | 34 (14.0) | 1 (0.9) | <0.001 |
| CKD | 21 (8.6) | 16 (13.8) | 0.14 |
| DM | 57 (23.5) | 36 (31.0) | 0.15 |
| Immunization status (vaccinated) | 142 (68.4) | 61 (52.6) | 0.30 |
| Days from symptoms onset (mean ±SD) | 3.8±2.7 | 3.8±2.8 | 0.89 |
| Days from positive PCR (mean ±SD) | 3.0±2.8 | 2.2±2.3 | 0.004 |
| Symptoms | 202 (84.9) | 102 (90.3) | 0.18 |
| Vital signs (mean±SD) | | | |
| Pulse (bpm) | 84.4±16.5 | 86.3±16.4 | 0.30 |
| Saturation (%) | 96.9±2.1 | 95.9±2.9 | <0.01 |
| Systolic Blood Pressure (mmHg) | 131.1±20.4 | 134.9±21.5 | 0.10 |
| Diastolic Blood Pressure (mmHg) | 73.7±11.4 | 76.4±14.3 | 0.05 |
| Body temperature (Celsius) | 37.3±0.78 | 37.4±0.82 | 0.68 |
| Laboratory results |  |  |  |
| WBC (K/uL) | 7.5±5.9 | 8.7±15.5 | 0.32 |
| Neutrophil/lymphocytes ratio | 5.5±6.6 | 4.7±5.0 | 0.31 |
| CRP (mg/dL) | 40.3±45.9 | 44.9±54.8 | 0.42 |
| CXR positive findings during presentation | 68 (41.2) | 47 (57.3) | 0.02 |
| Admitted to hospital | 84 (34.6) | 52 (44.8) | 0.04 |
| Severe disease outcome | 26 (10.7) | 24 (20.7) | 0.01 |
| 14-day desaturation | 24 (10.1) | 24 (20.7) | 0.008 |
| 45-day mortality | 4 (1.7) | 3 (2.6) | 0.68 |

Significantly more patients who were treated had a severe disease outcome (20.7% of the treated patients vs. 10.7% of those who were not, *P*=0.01). The patients who were treated with Casirivimab/Imdevimab were significantly older (62.2±18.3 years vs. 57.6±22.2 years, *P*=0.03), and had significantly higher rates of immunosuppressive disorders (48.3% vs 24.0%, *P*<0.001). Patients in the treated group had significantly lower room air saturation levels, albeit within the normal range (95.9%±2.9% vs. 96.9%±2.1%, *P*<0.01) and higher rates of abnormal (but not severe) findings on chest x-rays (57.3% vs. 41.2%, *P*=0.02).

### Patients who were treated with Casirivimab/Imdevimab

One-hundred and sixteen patients (52.6% females) were treated with Casirivimab/Imdevimab during their ED visit or hospital stay. That group’s median age was 64.5 years (IQR 51.0-75.7). The median time from symptom onset was 3 days (IQR 2-5 days) and 1 day from positive PCR results (IQR 1-3 days). Sixty-one patients (52.6%) were vaccinated at least twice with the COVID-19 vaccine, and 17 patients (14.7%) were vaccinated with a 3^rd^ dose. The median IgG level was 0.32 unit/ml (range: 0-151).

### Vaccinated vs. non-vaccinated patients and severe disease outcome among those who were treated with Casirivimab/Imdevimab

There was no significant difference in the occurrence of room air saturation <93% within 14 days of initial presentation or 45-day all-cause mortality between those who were vaccinated and those who were not (45.8% of the vaccinated patients vs. 54.3% of those who were not, *P*=0.49). An additional sub analysis failed to reveal any differences in the occurrence of a severe disease outcome between patients who received the 3^rd^ dose of vaccine and those who did not (12.5% and 15.2%, respectively, *P*>0.99).

### Antibody titers and severe disease outcome

There was no significant difference in the occurrence of room air saturation <93% within 14 days of initial presentation or 45-day all-cause mortality between the seronegative (IgG levels <1U/ml) and seropositive (IgG levels ≥1 U/ml) patients (24.1% vs. 17.1%, respectively, *P*=0.55) who were treated with Casirivimab/Imdevimab and whose antibodies levels had been documented. In addition, there was no significant correlation between antibody titers and severe disease outcome (Spearman correlation 0.04, *P*=0.73).

### Sub analysis of unvaccinated patients

Fifty-five of the 156 patients who were defined as unvaccinated (either were not vaccinated at all or were vaccinated with only 1 dose) were treated with Casirivimab/Imdevimab and 101 were not. There were no significant differences in the occurrence of severe disease outcome between patients who were treated with Casirivimab/Imdevimab and those who were not (23.6 % vs. 13.9 %, respectively, *P*=0.12).

### Sub analysis of vaccinated patients

Sixty-one of the 203 vaccinated patients were treated with Casirivimab/Imdevimab and 142 were not. There were no significant differences in the occurrence of severe disease outcome between the 2 groups (18.0% vs. 8.0%, respectively, *P*=0.56).

### Comparison of epidemiologic and clinical characteristics according to disease severity outcome

(Table 2). The patients with a severe disease outcome were significantly older than those whose disease outcome was not severe (68.7±19.6 years vs.57.5±20.9 years, respectively, *P*<0.001). There were significantly higher rates of patients with chronic kidney disease in the severe disease outcome group compared to the controls (24.0% vs. 8.1%, respectively, *P*<0.001). The mean number of days from symptom onset was significantly higher among patients with severe disease outcome (4.7±3.2 vs. 3.7±2.6, *P*=0.01). The mean room air saturation values were significantly lower for the severe disease outcome group (94.4±3.5 % vs. 96.9±2.0%, *P*<0.001), and the mean body temperature values were significantly higher (37.6±0.9°C vs. 37.3±0.7°C, *P*=0.008). The neutrophil/lymphocytes ratio and the CRP levels were significantly higher for the severe disease outcome group compared to the control group (7.0±7.6 vs. 4.9±5.8, *P*=0.03 and 73.7±57.0 mg/dl vs. 36.3±45.3 mg/dl, *P*<0.001, respectively). Finally, significantly more patients in the severe disease outcome group had abnormal chest x-ray findings at the presentation (77.8% vs. 39.6%, *P*<0.001). (Table 1).

**TABLE 2.** Comparison of epidemiological and clinical characteristics of patients with and without severe disease outcome

| Characteristic | No severe disease outcome (n=309) (N, %) | Severe disease outcome (n=50) (N, %) | P value |
| --- | --- | --- | --- |
| Age (mean±SD) | 57.5±20.9 | 68.7±19.6 | <0.001 |
| Sex (female) | 167 (53.8) | 23 (46.0) | 0.53 |
| Vaccination status |  |  |  |
| At least one dose | 183 (66.3) | 26 (55.3) | 0.18 |
| ≥2 doses | 180 (58.3) | 23 (46.0) | 0.12 |
| 3 doses | 51 (16.5) | 6 (12.0) | 0.53 |
| Recovered | 10 (3.7) | 2 (4.2) | >0.99 |
| Risk factors |  |  |  |
| Heart disease | 72 (23.3) | 17 (34.0) | 0.11 |
| Lung disease | 51 (16.6) | 8 (16.0) | >0.99 |
| Diabetes Mellitus | 74 (23.9) | 19 (38.0) | 0.05 |
| Chronic kidney disease | 25 (8.1) | 12 (24.0) | 0.002 |
| Immunosuppression | 100 (32.5) | 14 (28.0) | 0.62 |
| Pregnancy | 32 (10.4) | 3 (6.0) | 0.44 |
| Days from symptoms onset (mean±SD) | 3.7±2.6 | 4.7±3.2 | 0.01 |
| Days from positive PCR (mean±SD) | 2.7±2.6 | 3.0±2.9 | 0.40 |
| Symptoms | 266 (86.9) | 38 (84.4) | 0.64 |
| Vital signs (mean±SD) |  |  |  |
| Pulse (bpm) | 84.1±16.7 | 91.0±13.4 | 0.06 |
| Saturation (%) | 96.9±2.0 | 94.4±3.5 | <0.001 |
| Systolic Blood Pressure (mmHg) | 132.0±20.8 | 134.4±21.3 | 0.46 |
| Diastolic Blood Pressure (mmHg) | 74.8±12.4 | 73.4±13.1 | 0.48 |
| Body temperature (Celsius) | 37.3±0.7 | 37.6±0.9 | 0.008 |
| Laboratory results (mean±SD) |  |  |  |
| WBC (K/uL) | 7.6±9.2 | 9.5±13.8 | 0.23 |
| Neutrophil/lymphocytes ratio | 4.9±5.8 | 7.0±7.6 | 0.03 |
| CRP (mg/dL) | 36.3±45.3 | 73.7±57.0 | <0.001 |
| CXR positive findings at presentation | 80 (39.6) | 35 (77.8) | <0.001 |
| Casirivimab/Imdevimab treatment (yes) | 92 (29.8) | 24 (48.0) | 0.01 |
| Other treatment during hospital stay |  |  |  |
| Anticoagulation | 24 (7.8) | 31 (62.0) | <0.001 |
| Steroids | 39 (12.5) | 16 (34.8) | <0.001 |
| Abx | 20 (6.5) | 13 (26.0) | <0.001 |
| LOS (days) (mean±SD) | 3.7±6.3 | 6.6±6.7 | 0.01 |

### Predictors for severe disease outcome

Based on both the univariable analysis and the values reported in the current literature, factors with significance level of *P*<0.15 and those considered as either risk or protective factors for severe disease outcome were inserted into a multi-regression analysis to explore which ones were independent predictors for severe disease outcome. Due to the co-linearity between tachycardia and fever, we chose to insert body temperature into the model since it was more significant in the univariable analysis. In addition, due to a potential co-linearity between an abnormal finding in the chest x-ray and low saturation values (even within the normal range), we chose to insert room air saturation into the model since it is a relatively more accurate measurement. Chronic kidney disease (aOR=3.51 [95% CI: 1.34-9.20], *P*=0.01), lower saturation (aOR=0.7 [95%

CI: 0.58–0.85], *P*<0.01), and higher CRP levels (aOR=1.01 [95% CI: 1.00 – 1.01], *P*=0.008) emerged as independent risk factors of severe disease outcome in the multi-regression model. Positive immunization status was found to be an independent protective factor of severe disease outcome (aOR=0.33 [95% CI: 0.14–0.77], *P*=0.01). Casirivimab/Imdevimab treatment was not found to be an independent protective factor for severe disease outcome on a multivariable logistic regression model (aOR=1.54 [95% CI: 0.71–3.34], *P*=0.26) (Table 3).

**TABLE 3.**
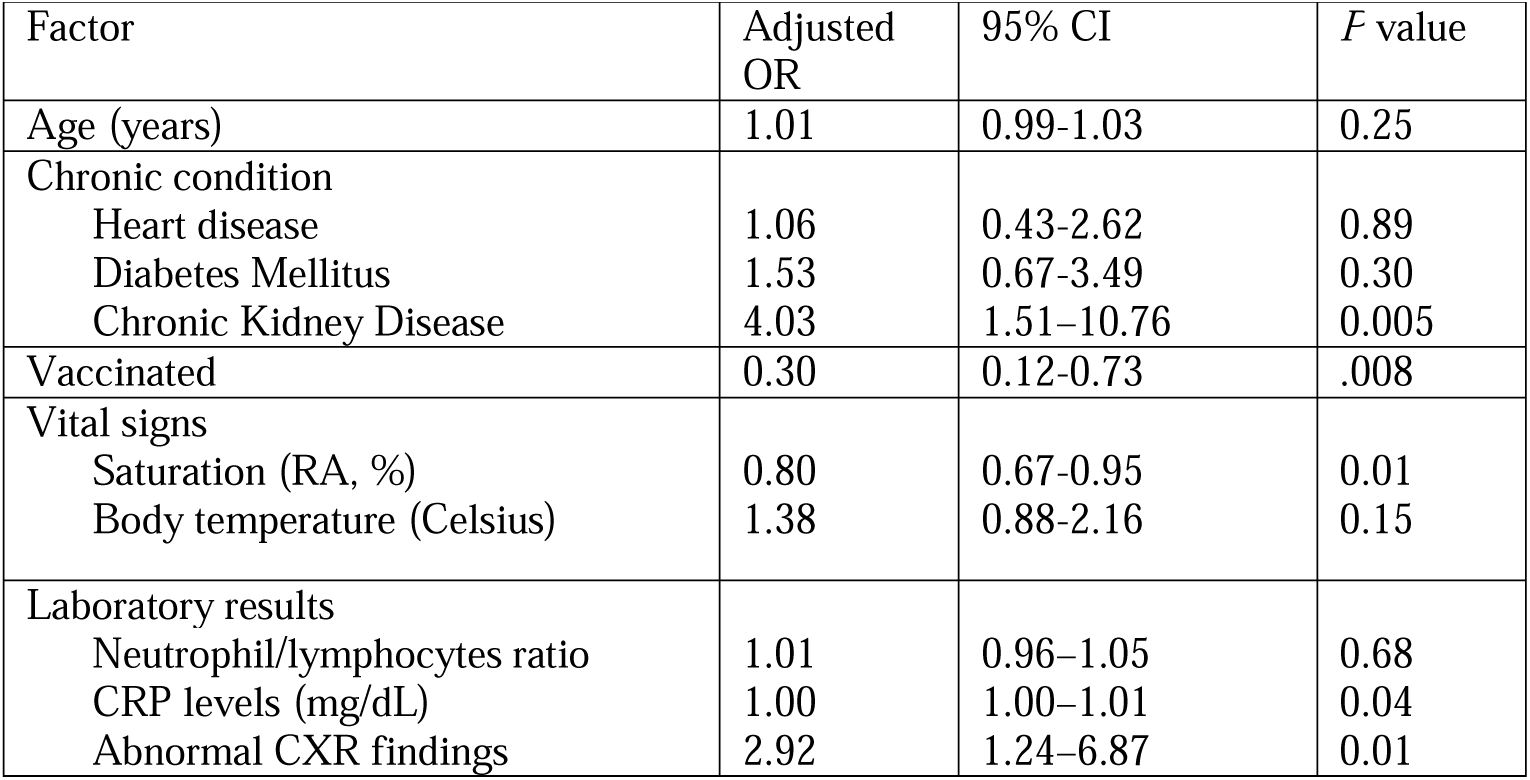
Multivariable logistic regression model for severe disease outcome

| Factor | Adjusted OR | 95% CI | *P* value |
| --- | --- | --- | --- |
| Age (years) | 1.01 | 0.99-1.03 | 0.25 |
| Chronic condition |  |  |  |
| Heart disease | 1.06 | 0.43-2.62 | 0.89 |
| Diabetes Mellitus | 1.53 | 0.67-3.49 | 0.30 |
| Chronic Kidney Disease | 4.03 | 1.51–10.76 | 0.005 |
| Vaccinated | 0.30 | 0.12-0.73 | .008 |
| Vital signs |  |  |  |
| Saturation (RA, %) | 0.80 | 0.67-0.95 | 0.01 |
| Body temperature (Celsius) | 1.38 | 0.88-2.16 | 0.15 |
| Laboratory results |  |  |  |
| Neutrophil/lymphocytes ratio | 1.01 | 0.96–1.05 | 0.68 |
| CRP levels (mg/dL) | 1.00 | 1.00–1.01 | 0.04 |
| Abnormal CXR findings | 2.92 | 1.24–6.87 | 0.01 |

## DISCUSSION

The aim of this study was to assess the “real-life” contribution of Casirivimab/Imdevimab monoclonal antibody therapy in preventing severe disease outcome in high-risk patients with SARS-CoV-2 Delta variant infection. In contrast to previous reports (18), we found that Casirivimab/Imdevimab did not significantly affect the 45-day mortality rates or the 14-day air room desaturation levels, nor was it found to be an independent protective factor against severe disease outcome. Moreover, it had a significant association with severe disease outcome in the univariable analysis.

Several factors may explain our observation. First, our sub-analysis revealed that patients who were treated with Casirivimab/Imdevimab had higher rates of immunosuppressive conditions, which had been reported to be associated with a higher likelihood of hospitalization following monoclonal antibody treatment (19). In addition, we found, in consistent with a recent report (20), that a positive immunization status, with at least 2 doses of vaccine, was an independent protective factor against severe disease outcome. However, the COVID-19 mRNA vaccine had lower vaccine effectiveness among immunocompromised patients compared to the immunocompetent controls (21), and studies found that the ability of the former to develop high neutralizing antibody titers and to be protected against severe COVID-19 outcomes were limited compared to the latter (22, 23). These observations may partially explain the higher rates of severe outcome among patients who received Casirivimab/Imdevimab treatment, compared to those who were not, in our cohort.

We found that patients who were treated with Casirivimab/Imdevimab had significantly lower room air saturation levels (within the normal ranges), higher rates of abnormal chest x-ray findings (already at their initial presentation), and higher rates of hospital admissions. These associations may represent a selection bias, since clinicians may chose the more severely ill patients within the high-risk group for treatment with Casirivimab/Imdevimab. Wienriech et al (24) found that there was a steeper reduction in the viral load among patients treated with Casirivimab/Imdevimab who were seronegative at the time of diagnosis compared to those who were seropositive. In our study, more than one-half of the Casirivimab/Imdevimab recipients were vaccinated with at least 2 COVID-19 vaccine doses and were seropositive at some level. This could explain, at least in part, why Casirivimab/Imdevimab treatment had limited additive value (24). Our sub-analysis in which we compared unvaccinated with vaccinated patients showed no differences in the occurrence of severe disease outcome between those who underwent Casirivimab/Imdevimab therapy and those who did not. Moreover, there was no significant difference in the occurrence of severe disease outcome between seropositive and seronegative patients.

We, as others (19,25), found chronic kidney disease to be an independent risk factor for severe disease outcome. The high prevalence of comorbidities in patients with chronic kidney disease, such as hypertension, cardiovascular disease, and diabetes mellitus, might contribute to the poorer outcomes among those COVID-19 patients.

Greater disease severity was found to be associated with older age in a series of multivariable-adjusted analyses based upon COVID-19 patient cohorts (26). Although age was significantly higher in our severe disease outcome group in the univariable analysis, it was not an independent predictor for severe disease outcome in the multi-regression model, potentially due to co-factors such as chronic diseases. It should be borne in mind that our patients were selected by either older age or chronic disease and that our cohort already represents a high-risk group for severe disease outcome, and that our findings should be interpreted accordingly.

It is well documented (27-29) that room air desaturation at presentation is a risk factor for severe disease, and we found lower saturation levels at ED presentation - even those within normal range - to be an independent risk factor for severe COVID-19 outcome. In addition, our findings of an association between abnormal x-ray findings with severe disease outcome is consistent with those of previous studies (27,30). Our demonstration that CRP levels were an independent predictor for severe disease outcome is consistent with other studies (25,30), which demonstrated that high levels of serum CRP are key markers of disease progression and a risk factor for mortality of COVID-19 patients with severe disease.

The treatment with Casrivimab/Imdevinab, as with other mAbs, has the potential to lead to the development of resistant variants (28). The Omicron (B.1.1.529) SARS-CoV-2 is quickly becoming the dominant variant (29), and it reportedly has some immune evasion against currently used mAbs (32). A recent study found that Casrivimab/Imdevinab lost its antiviral activity against the Omicron variant (33). These data, taken together with our results, raise some doubt about the benefit of Casrivimab/Imdevinab for treating new SARS-CoV-2 variants.

Our study has several limitations related to its retrospective nature. One is that some relevant data documented in the medical charts may have been incomplete, such as body mass index values and documented SARS-CoV-2 IgG levels. In addition, it was difficult to retrospectively discern how the co–occurrence of COVID-19 infection was affecting the clinical course of patients who were diagnosed with COVID-19 during their hospitalization for other reasons.

## CONCLUSION

We found no added benefit to the administration of Casrivimab/Imdevinab monoclonal antibody therapy to a mostly vaccinated high-risk population with an early delta variant of SARS-COVID-19 infection. Additional studies of new variants in the vaccination era are needed to explore the effect of monoclonal antibody therapy on the severity of disease outcome.

## Data Availability

All data produced in the present study are available upon reasonable request to the authors

